# Amyloid deposition in granuloma of tuberculosis patients: A pilot study

**DOI:** 10.1101/2021.02.08.21250526

**Authors:** Shreya Ghosh, Chayanika Kala, Akansha Garg, Ashwani Kumar Thakur

**Author notes:** Corresponding author: Dr. Ashwani K Thakur, Department of Biological Sciences and Bioengineering, Mehta Family Center for Engineering in Medicine, Indian Institute of Technology Kanpur, Uttar Pradesh-208016, India. **E-mail:**.

## Abstract

The formation of granuloma is one of the characteristic features of tuberculosis. Besides, rise in the concentration of serum amyloid A (SAA) protein is the indicator for chronic inflammation associated with tuberculosis. The predisposition of SAA driven secondary amyloidosis in tuberculosis is well documented. However, SAA derived amyloid onset and deposition start sites are not well chracterised in tuberculosis and other chronic inflammatory conditions. We hypothesized that granuloma could be a potential site for amyloid deposition because of the presence of SAA protein and proteases that cleave SAA into aggregation prone fragments. 150 tuberculosis patients were screened and biopsies were collected from the affected organs of these patients. 20 patients showed eosinophilic hyaline rich deposits within and surrounding granuloma respectively. They were further screened for the presence of amyloid deposits. The hyaline material, upon Congo red staining exhibited characteristic apple green birefringence under polarized light, confirming deposition of amyloid. Further upon immuno histochemical staining with anti-SAA antibody, the amyloid enriched areas showed positive immunoreactivity. In this pilot study, wexx have shown granuloma as a potential site for serum amyloid A derived amyloid deposition in tuberculosis patients. This study would set a stage to expxand the clinical and fundamental research for understanding the mechanism of amyloid formation in granuloma underlying tuberculosis and other chronic inflammatory conditions.

## 1. Introduction

Tuberculosis (TB) is one of the leading airborne disease, caused by *Mycobacterium tuberculosis* infection [1]. TB is highly prevalent across the world, affecting 10 million people in a year [2]. Amongst all WHO regions in South-East Asia, India is reported as the highest TB burden country accounting for 26% of the global TB prevalence in the year 2019 [2].

Lungs are the most common site for mycobacterial infection in majority of cases. However, organs systems like gastrointestinal, musculoskeletal, lymphoreticular, skin, liver, reproductive and the central nervous system are also affected by tuberculosis [3]. Despite, of having various treatment options, it remains one of the leading causes of morbidity. Existence of several multidrug resistant mycobacterial tuberculosis strains could be one of the reasons behind it [4].

During the chronic infections including tuberculosis, serum amyloid A (SAA) levels often increases to 1000 folds [5]. This induces cleavage of the SAA protein by cathepsins (cysteine proteases) and MMPs to form N-terminal fragment [6,7]. This truncated form of SAA as well as full-length protein gets deposited as amyloid fibrils in the extracellular spaces of various organs especially in liver and spleen [8-10]. Besides, presence of proteases like MMPs and Cathepsins have been reported in AA amyloid deposits, suggesting their putative roles in amyloid formation [7,11,12]. SAA was first identified as the “amyloid of unknown origin” underlying chronic inflammation in 1961 by Benditt and Eriksen [13]. In the subsequent years other studies depicted the complete sequence of protein as well as the amyloidogenic N-terminal fragment [14-16]. Later it was named as serum amyloid A protein because it was the first non-immunoglobulin serum protein identified to have a role in amyloid formation [5].

Prevalence of secondary amyloidosis underlying chronic inflammatory conditions in tuberculosis patients has been reported since early 1900s. In a study carried out at Uppsala University with 65-83 years old formalin fixed organs from pulmonary TB patients, 48 out of 156 organ autopsies was reported to be associated with amyloidosis [17,18]. The incidence of AA amyloidosis is quite higher in developing countries including India [19]. Various epidemiological studies on Indian population have revealed secondary amyloidosis as one of the major outcomes of pulmonary tuberculosis [20-24].

One of the major signature of tuberculosis patients is formation of granuloma, comprising of aggregate of epitheloid cells, mature macrophages and Langhans giant cells [25]. It is formed as a result of first line of host defence to eradicate the microbes. But in later stages, it is utilized by the mycobacterial species as a vehicle for its cellular expansion within host system [26]. MMPs play a vital role in tuberculous granuloma formation and disease progression post Mtb infection [27]. Enhanced expression of MMP 9, MMP 1 and MMP 3 is often seen within tuberculous granuloma whereas MMP 7 expression is mostly seen at its periphery [28-31]. Additionally, in a rabbit model for pulmonary tuberculosis, enhanced expression of cathepsins has been observed in the granuloma [32]. Similarly, tuberculosis patient derived tissues showed enhanced expression of cathepsins B and K in the granulomatous structure [33,34].

SAA is known to have a high affinity for the macrophages [35]. Chen et.al in 2010 reported that SAA expression was localized at the periphery of tuberculous granuloma [36]. As SAA, after proteolytic cleavage drives the systemic form of amyloidosis in tuberculosis patients, we hypothesised that the presence of SAA, MMPs and cathepsins might account for the onset of amyloid formation and deposition in the granulomas. In this study, we have shown the presence of SAA derived amyloid deposits in the granuloma of tuberculosis patients.

Congo red (CR) staining is considered as the gold standard for identifying amyloid deposits in the affected tissues [37,38]. The characteristic apple green birefringence under polarized light along with its transformation from apple green to reddish orange or bluish green (based on fibril orientation), upon changing the angle of polarizer is a strong signature of amyloid fibrils [39,40]. In this study we have utilized this birefringence property of CR staining to trace the amyloid deposits in tuberculous granuloma.

## 2. Methods

### 2.1 Sample collection

In a two-year single center study carried out from 2018-2020, 150 patients were screened positive for tuberculosis based on sputum examination, histological features and other distinct biochemical parameters in blood. Tissue specimens after biopsy of the respective affected organs of these TB patients were then collected after taking their informed consent. The ethics approval (**Ref No**: EC/BMHR/2021/19 and IITK/IEC/2021-22/I/5) for this study was obtained from the human ethics committee of both GSVM medical college and Indian Institute of Technology Kanpur respectively.

### 2.2 Histological examination of the tissue specimens

A series of staining procedures were carried out for the identification of structural features and presence of amyloid deposits in the FFPE tissue sections. The detailed protocols are outlined in the following paragraphs.

#### 2.2.1 Hematoxylin-Eosin staining

Deparaffinized and rehydrated sections were put in Hematoxylin-Mayer’s solution for 2 minutes followed by a brief wash in distilled water. It was then counterstained with 1% eosin solution followed by subsequent dehydration and xylene treatment. The slides were then mounted in mounting media and observed in bright light under a microscope.

#### 2.2.2 Thioflavin T staining

The FFPE sections were deparaffinized and rehydrated by different grades of ethanol. Hydrated sections were then stained with 1% Thioflavin T (Sigma Aldrich, Catalog no: T-3516) solution for 2 minutes. Excess stain was removed by rinsing the slide containing tissue specimen in two changes of water, not exceeding 10 seconds for each. Finally it was dehydrated in two changes of 100% ethanol and xylene. Finally, the stained section was mounted in hydrophobic mounting media and was observed under a fluorescence microscope in blue light.

#### 2.2.3 Congo red Staining

Stock and working solutions were prepared according to the previously published protocol [41]. Sections were deparaffinized, rehydrated and stained with Hematoxylin-Mayer’s solution followed by brief rinsing in water to remove excess stain. It was then transferred to solution A for 20 minutes immediately followed by 20 minutes incubation in solution B. Sections were then rinsed briefly in two changes of absolute ethanol, not exceeding for 10 seconds each followed by xylene treatment. It was then mounted in synthetic mounting media. Stained sections were then visualized using a filter emitting green light and both crossed and uncrossed polariser respectively under a microscope.

#### 2.2.4 Immunohistochemistry

Sections were deparaffinized and rehydrated followed by heat induced antigen retrieval in 10 mM sodium citrate buffer containing 0.1% tween 20. It was then treated with 3% hydrogen peroxide to inhibit endogenous peroxidase activity. Sections were then incubated overnight with primary antibody against SAA protein (Santa crutz, Catalog no: sc-59679) in 1:100 dilution followed by secondary antibody (Santa crutz, Catalog no: sc-516102) in 1:50 dilution. Sections were developed using DAB substrate (Sigma Aldrich, Catalog No: D 3939). Developed sections were then dehydrated, mounted in mounting media and visualized under a microscope in bright light.

## 3. Results and discussion

Histopathological diagnosis of all the 150 TB patients showed that the majority of cases were having caseous necrosis (80/150) followed by non-necrotizing granulomatous inflammation (10/150), non-caseating granulomatous inflammation (30/150), sinus hyperplasia (20/150), follicular hyperplasia (5/150) and fibrosis (5/150) (Figure 1). 25 out of 150 TB patients (Table 1) showed presence of hyaline rich eosinophilic deposits in the tissue specimens upon H& E staining. But the granulomatous lesions in 20 patients was observed to be laden with this hyaline material (Figure 1 A, B). Amyloid has been reported to show amorphous hyaline like appearance since ages [42]. Hence, these patients were further screened for tracing the presence of amyloid in this study.

**Table 1:** Preliminary information of the TB patients suspected of amyloidosis

| Characteristics | Number |
| --- | --- |
| Total subjects in this study (male , female) | 20 (13, 7) |
| Mean age | 28 ± 9.32 |
| **Primary diagnosis** |  |
| Tuberculous lymphadenitis | 8 |
| Tuberculous enteritis | 5 |
| Tuberculous endometritis | 1 |
| Cutaneous tuberculosis | 1 |
| Tubercular ileitis | 2 |
| Tuberculous appendicitis | 1 |
| Tubercular oophoritis | 1 |
| Foot osteoarticular tuberculosis | 1 |
| Tuberculous synovitis | 1 |

**Figure 1:**
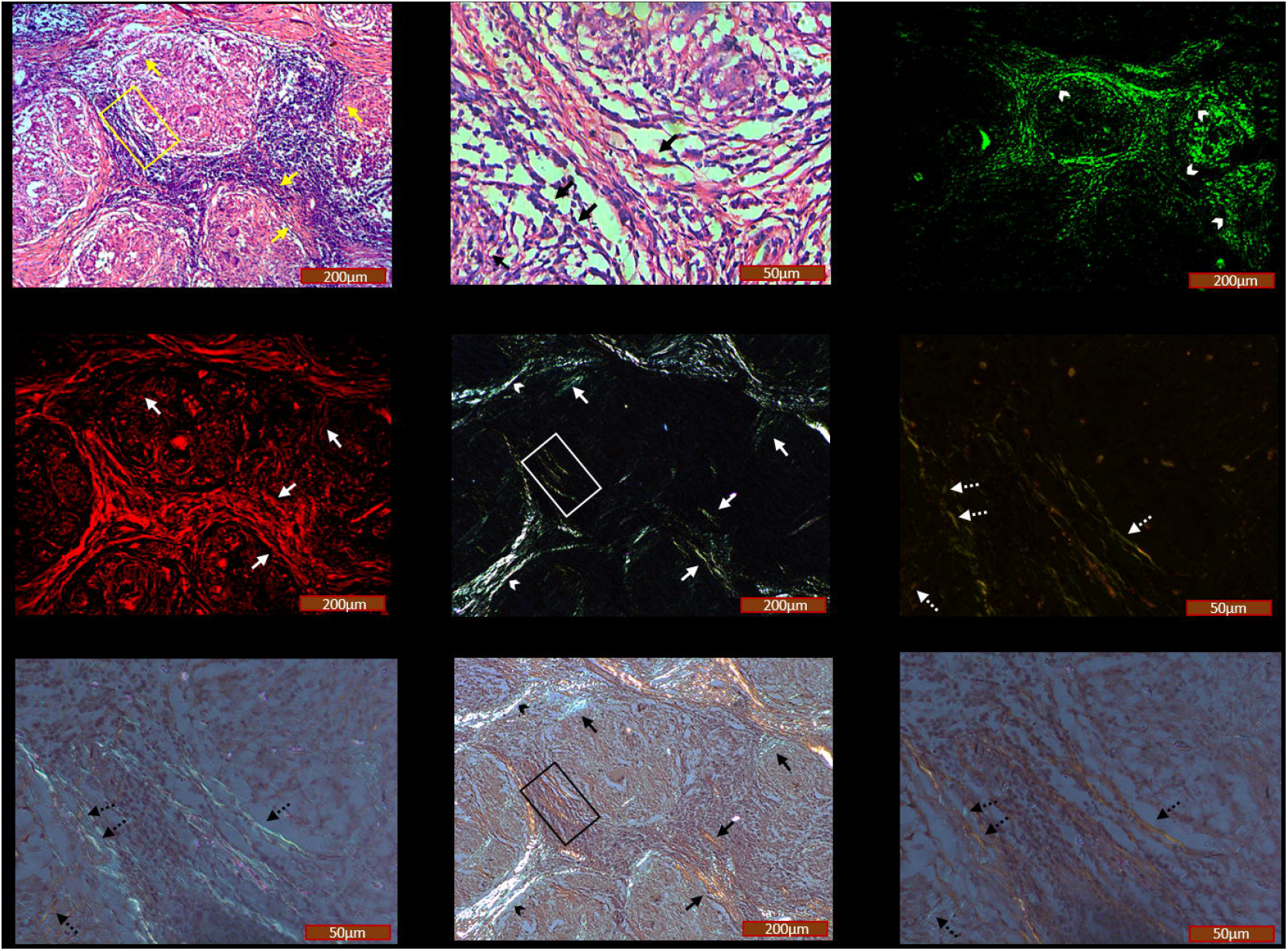
Histology representation of the affected Synovium tissue specimen of one of the tuberculosis patients, suspected of amyloidosis. Eosinophilic hyaline rich deposits were mostly seen around the periphery of the granulomatous structure (indicated by yellow solid arrows and marked by black solid arrows in the magnified image of yellow square boxed area) (A, B). Upon staining the same section with Thioflavin T, an amyloid specific probe, green fluorescence was observed corresponding to the areas of hyaline material deposits (indicated by white arrow heads) (C). Further, Congo red staining of the same tissue section showed red fluorescence (indicated by solid arrows) (D) along with apple green birefringence (indicated by white solid arrows (E) and marked by white dashed arrows in the magnified image (F) of white square boxed area in E), consistent with the areas having eosinophilic deposits. A colour transition, depending on the orientation of fibrils was observed upon rotating the polarizer by 10^0^ either in clockwise or anticlockwise direction, confirming the presence of amyloid deposits (indicated by black solid arrows (H) and marked by black dashed arrows in the magnified images (G, I) of black square boxed area in H). However, this colour transition was not observed for the collagen that remained bluish white irrespective of the specimen kept in either crossed or uncrossed polarisers (indicated by white arrow heads in E and black arrow heads in H respectively). **Scale bar**: 200 μm. **Scale bar for magnified images (B, F, G, I)**: 50 μm.

Haematological parameters like WBC count, platelet count, RBC count, haemoglobin and erythrocyte sedimentation rate are often seen altered in tuberculosis patients [43-46]. In our study, the blood profile of all the TB patients including those suspected for amyloidosis (Table 2) also reflected similar alteration in these inflammatory markers. It is well known that tuberculosis predisposes systemic secondary amyloidosis, affecting normal functioning of kidney [47]. Proteinuria and Hypoalbuminea are the two major clinical manifestations of amyloid associated kidney dysfunction [48]. However, the renal function test (RFT) profile (Table 3) and 24 hour urine excretory protein level of these patients were normal. This implied that there was deposition of eosinophilic hyaline material in the granuloma of the affected tissue in these patients without renal involvement.

**Table 2:**
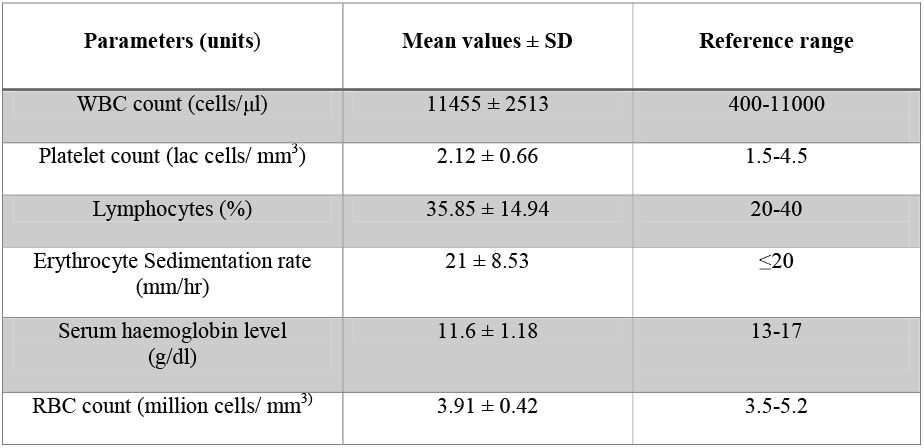
Blood profile of the amyloid suspected tuberculosis patients

**Table 3:** RFT profile of the amyloid suspected tuberculosis patients

| Parameters (units) | Mean values ± SD | Reference range |
| --- | --- | --- |
| Serum creatinine (mg/dl) | 1.06 ± 0.13 | 0.5-1.5 |
| Serum albumin (g/dl) | 4.5 ± 0.48 | 3.5-5.2 |
| Serum urea (mg/dl) | 30 ± 8.16 | 10-45 |
| Ionic calcium (mg/dl) | 5.04 ± 0.17 | 4.5-5.5 |

SAA, a known amyloidogenic protein is reported to be localized in the periphery of tuberculous granuloma [36]. Besides, it is well known that MMPs and cathepsins, a class of endopeptidase that cleaves SAA into amyloidogenic fragments are strongly expressed in the tuberculous granuloma [28-32,49]. Altogether, it heightened our suspicion for amyloid presence in the granuloma. Initially, these biopsies were stained with an amyloid detecting molecular rotor, Thioflavin T [38] showing a bright green fluorescence in and around the granuloma of these 20 patients (Figure 1C). Further, to confirm our observation, the gold standard Congo red staining was done in the biopsies of these patients to identify amyloid deposits. The granulomatous structures were observed to exhibit characteristic red fluorescence (Figure 1 D) and apple green birefringence under polarized light respectively (Figure 1 E, F) both within and around its periphery. Upon changing the angle of the polariser by 10^0^ either in clockwise or anticlockwise direction, a transition from apple green birefringence to either reddish orange coloration or bluish green was also observed, depending on the orientation of the fibrils (Figure 1 G, H, I). Pathologists use this characteristic feature for confirming amyloid presence and reduce the ambiguity [39,40]. Hence, it confirmed the presence of amyloid deposits in these patients. Immunohistochemistry of these sections using antibody against serum amyloid A protein showed patchy expression of SAA protein throughout the granuloma. Additionally, presence of SAA was also observed in the amyloid enriched areas (Figure 2), suggesting granuloma as the potential site for amyloid deposition in active tuberculosis patients.

**Figure 2:**
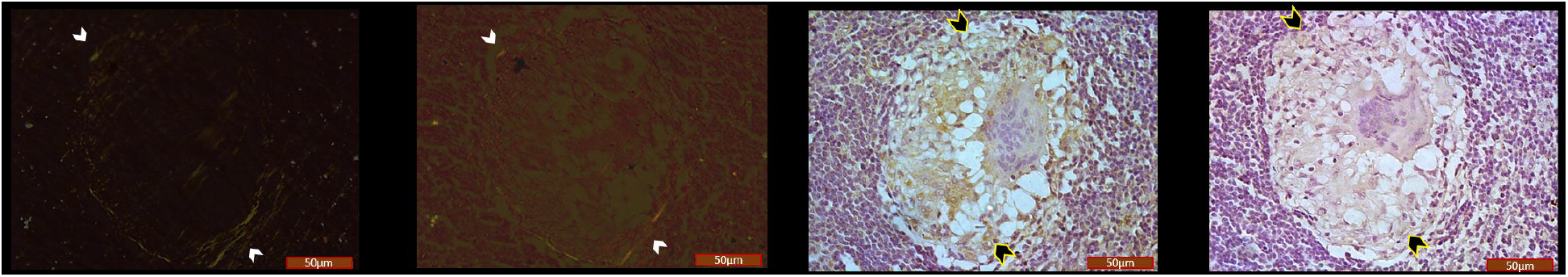
Representative microscopic images of the affected lymph node section of one of the 20 TB patients showing presence of serum amyloid A protein in patches in the granulomatous structure as well as in the amyloid enriched areas respectively (C) (indicated by white arrow heads in A, B and yellow outlined black arrow heads in C respectively). But the antibody control (D) showed no positive signal for serum amyloid A protein indicating that the amyloid deposits are AA derived. **Scale bar**: 50μm

Serum amyloid A protein is reported to act as a bridge between mycobacterial infection and granulomatous inflammatory response [36]. In case of persistent inflammation underlying mycobacterial infection, liver produces increased amount of SAA protein to combat bacterial action through mediated immune response [50]. In sarcoidosis, Chen *et al*. predicted presence of serum amyloid A protein in the granuloma in association with other extracellular matrix proteins [36]. Additionally, proteases like MMPs and cathepsins have been recently reported to be up regulated in tuberculous granuloma [51]. Considering these facts, we assume that the systemic serum amyloid A protein after getting cleaved by these proteases becomes amyloidogenic. This in turn might seed the SAA protein already present in the granulomatous structure, resulting in amyloid deposition. Several proteins of *mycobacterium tuberculosis* has been reported to amyloid like fibrils [52-55]. Besides, activated macrophages, one of the major granuloma components have been known to promote fibrillation of serum amyloid A both *in-vitro* and *in-vivo* systems [56-58]. Altogether, it is anticipated that apart from seeding, synergistic interactions between the granuloma components and serum amyloid A protein might also account for the onset of amyloid formation and deposition in tuberculous granuloma (Figure 3).

**Figure 3:**
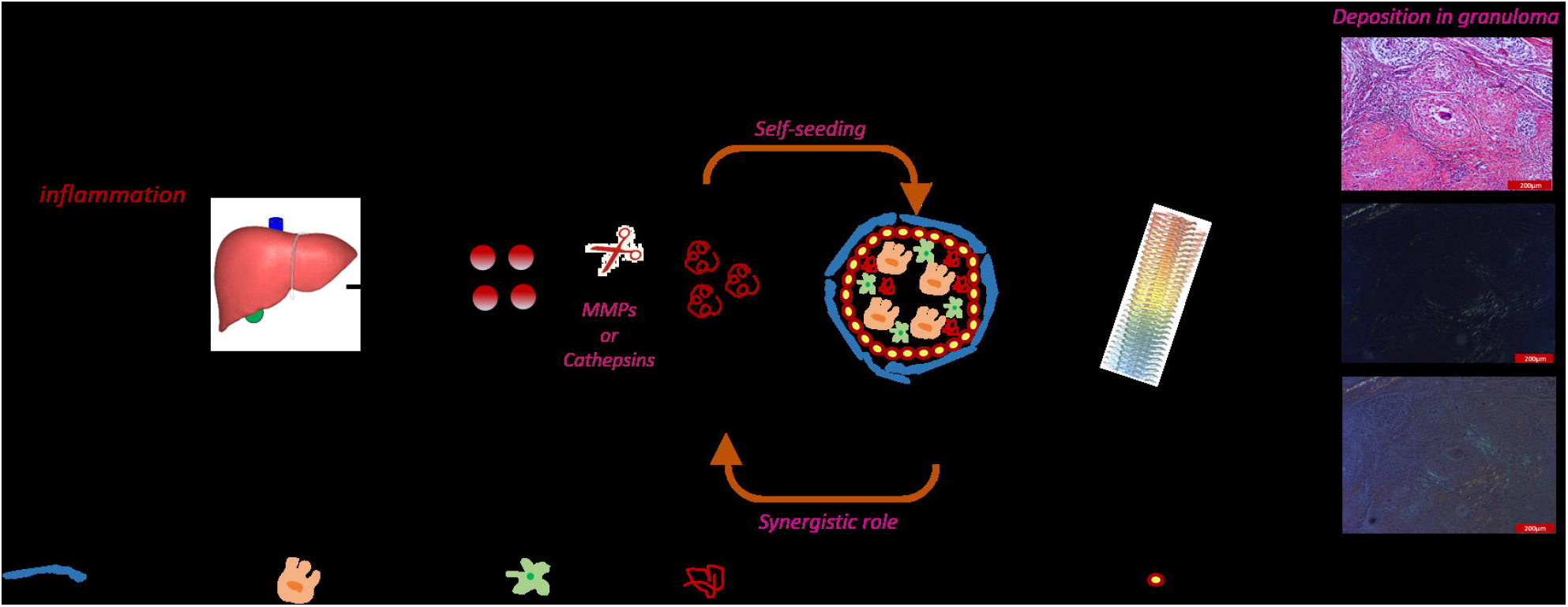
Probable mechanism underlying Serum amyloid A derived amyloid formation in tuberculous granuloma.

## Conclusion

To the best of our knowledge, this is the first report showing presence of SAA derived amyloid deposits in tuberculous granuloma. These findings have paved new ways for exploring either the synergistic or independent role of granuloma components and SAA in driving amyloid formation in tuberculosis patients. Based on these findings, mechanism of amyloid formation can be traced in various stages of granuloma development using various animal models [59,60]. The clinical manifestations of amyloidosis are generally identified in the later stages of tuberculosis. Hence, it remains undiagnosed and untreated in the initial stages of tuberculosis patients. This study would set a platform for the clinicians to diagnose amyloid onset and progression even in early stages of tuberculosis. This in turn would enable the clinicians to provide targeted therapy to tuberculosis patients having amyloidosis.

## Data Availability

All data produced in the present work are contained in the manuscript

## Abbreviations

TB: tuberculosis
WHO: world health organization
FDA: Food and Drug Administration
SAA: serum amyloid A
H&E: Hematoxylin and eosin
FFPE: formalin fixed paraffin embedded
CR: Congo red
MMPs: matrix metalloproteinases
mM: milli molar
Mtb: *Mycobacterium tuberculosis*
SD: standard deviation

## References

1. Furin J, Cox H, Pai M. Tuberculosis. The Lancet 2019; 393 (10181):1642–1656.

2. Organization WH. Global tuberculosis report 2020. Global tuberculosis report 20202020.

3. Loddenkemper R, Lipman M, Zumla A. Clinical Aspects of Adult Tuberculosis. Cold Spring Harb Perspect Med 2015; 6 (1):a017848–a017848.

4. Seung KJ, Keshavjee S, Rich ML. Multidrug-Resistant Tuberculosis and Extensively Drug-Resistant Tuberculosis. Cold Spring Harb Perspect Med 2015; 5 (9):a017863–a017863.

5. Sack GH. Serum amyloid A – a review. Molecular Medicine 2018; 24 (1):46.

6. Stix B, Kähne T, Sletten K, Raynes J, Roessner A, Röcken C. Proteolysis of AA amyloid fibril proteins by matrix metalloproteinases-1, -2, and -3. Am J Pathol 2001; 159 (2):561–570.

7. Röcken C, Menard R, Bühling F, Vöckler S, Raynes J, Stix B, et al. Proteolysis of serum amyloid A and AA amyloid proteins by cysteine proteases: cathepsin B generates AA amyloid proteins and cathepsin L may prevent their formation. Annals of the Rheumatic Diseases 2005; 64 (6):808–815.

8. Lu J, Yu Y, Zhu I, Cheng Y, Sun PD. Structural mechanism of serum amyloid A-mediated inflammatory amyloidosis. Proceedings of the National Academy of Sciences 2014; 111 (14):5189–5194.

9. Westermark GT, Fändrich M, Westermark P. AA Amyloidosis: Pathogenesis and Targeted Therapy. Annual Review of Pathology: Mechanisms of Disease 2015; 10 (1):321–344.

10. Real de Asúa D, Costa R, Galván JM, Filigheddu MT, Trujillo D, Cadiñanos J. Systemic AA amyloidosis: epidemiology, diagnosis, and management. Clin Epidemiol 2014; 6:369–377.

11. Muller D, Roessner A, Rocken C. Distribution pattern of matrix metalloproteinases 1, 2, 3, and 9, tissue inhibitors of matrix metalloproteinases 1 and 2, and α2-macroglobulin in cases of generalized AA-and AL amyloidosis. Virchows Archiv 2000; 437 (5):521–527.

12. Röcken C, Stix B, Brömme D, Ansorge S, Roessner A, Bühling F. A putative role for cathepsin K in degradation of AA and AL amyloidosis. The American journal of pathology 2001; 158 (3):1029–1038.

13. Benditt EP, Eriksen N. AMYLOID. II. STARCH GEL ELECTROPHORETIC ANALYSIS OF SOME PROTEINS EXTRACTED FROM AMYLOID. Archives of pathology 1964; 78:325–330.

14. Benditt EP, Eriksen N, Hermodson MA, Ericsson LH. The major proteins of human and monkey amyloid substance: Common properties including unusual N-terminal amino acid sequences. FEBS letters 1971; 19 (2):169–173.

15. Levin M, Franklin EC, Frangione B, Pras M. The amino acid sequence of a major nonimmunoglobulin component of some amyloid fibrils. The Journal of clinical investigation 1972; 51 (10):2773–2776.

16. Ein D, Kimura S, Glenner GG. An amyloid fibril protein of unknown origin: partial amino-acid sequence analysis. Biochemical and biophysical research communications 1972; 46 (2):498–500.

17. Westermark P, Nilsson GT. Demonstration of amyloid protein AA in old museum specimens. Archives of pathology & laboratory medicine 1984; 108 (3):217–219.

18. Westermark GT, Fändrich M, Westermark P. AA amyloidosis: pathogenesis and targeted therapy. Annual review of pathology 2015; 10:321–344.

19. Nienhuis HLA, Bijzet J, Hazenberg BPC. The Prevalence and Management of Systemic Amyloidosis in Western Countries. Kidney Dis (Basel) 2016; 2 (1):10–19.

20. Chugh K, Datta B, Singhal P, Jain S, Sakhuja V, Dash SJPmj. Pattern of renal amyloidosis in Indian patients. 1981; 57 (663):31–35.

21. Mehta HJ, Talwalkar NC, Merchant MR, Mittal BV, Suratkal LH, Almeida AF, et al. Pattern of renal amyloidosis in western India. A study of 104 cases. The Journal of the Association of Physicians of India 1990; 38 (6):407–410.

22. Dixit R, Gupta R, Dave L, Prasad N, Sharma S. Clinical profile of patients having pulmonary tuberculosis and renal amyloidosis. Lung India 2009; 26 (2):41–45.

23. Engineer D, Kute V, Patel H, Shah P. Clinical and laboratory profile of renal amyloidosis: A single-center experience. Saudi Journal of Kidney Diseases and Transplantation 2018; 29 (5):1065–1072.

24. Sharma S, Mathur M, Prasad D, Singh AP, Garsa R, Kumar R, et al. Are we Treating or Curing Tuberculosisã Profile of Secondary Renal Amyloidosis in Patients Receiving Anti Tubercular Treatment. BANTAO Journal 2014; 12 (2):103–107.

25. Petersen HJ, Smith AM. The role of the innate immune system in granulomatous disorders. Front Immunol 2013; 4:120–120.

26. Pagán AJ, Ramakrishnan L. Immunity and Immunopathology in the Tuberculous Granuloma. Cold Spring Harb Perspect Med 2014; 5 (9):a018499.

27. Sabir N, Hussain T, Mangi MH, Zhao D, Zhou X. Matrix metalloproteinases: Expression, regulation and role in the immunopathology of tuberculosis. Cell Proliferation 2019; 52 (4):e12649.

28. Price NM, Gilman RH, Uddin J, Recavarren S, Friedland JS. Unopposed Matrix Metalloproteinase-9 Expression in Human Tuberculous Granuloma and the Role of TNF-α-Dependent Monocyte Networks. The Journal of Immunology 2003; 171 (10):5579–5586.

29. Elkington PT, Nuttall RK, Boyle JJ, O’Kane CM, Horncastle DE, Edwards DR, et al. Mycobacterium tuberculosis, but not vaccine BCG, specifically upregulates matrix metalloproteinase-1. American journal of respiratory and critical care medicine 2005; 172 (12):1596–1604.

30. Green JA, Elkington PT, Pennington CJ, Roncaroli F, Dholakia S, Moores RC, et al. Mycobacterium tuberculosis upregulates microglial matrix metalloproteinase-1 and-3 expression and secretion via NF-κB–and activator protein-1–dependent monocyte networks. The Journal of Immunology 2010; 184 (11):6492–6503.

31. Gupta RK, Haris M, Husain N, Saksena S, Husain M, Rathore RK. DTI derived indices correlate with immunohistochemistry obtained matrix metalloproteinase (MMP-9) expression in cellular fraction of brain tuberculoma. Journal of the neurological sciences 2008; 275 (1-2):78–85.

32. Kubler A, Larsson C, Luna B, Andrade BB, Amaral EP, Urbanowski M, et al. Cathepsin K Contributes to Cavitation and Collagen Turnover in Pulmonary Tuberculosis. The Journal of Infectious Diseases 2015; 213 (4):618–627.

33. Bühling F, Reisenauer A, Gerber A, Krüger S, Weber E, Brömme D, et al. Cathepsin K--a marker of macrophage differentiation? The Journal of pathology 2001; 195 (3):375–382.

34. Chai Q, Lu Z, Liu Z, Zhong Y, Zhang F, Qiu C, et al. Lung gene expression signatures suggest pathogenic links and molecular markers for pulmonary tuberculosis, adenocarcinoma and sarcoidosis. Communications Biology 2020; 3 (1):604.

35. Kisilevsky R, Subrahmanyan L. Serum amyloid A changes high density lipoprotein’s cellular affinity. A clue to serum amyloid A’s principal function. Laboratory investigation; a journal of technical methods and pathology 1992; 66 (6):778–785.

36. Chen ES, Song Z, Willett MH, Heine S, Yung RC, Liu MC, et al. Serum amyloid A regulates granulomatous inflammation in sarcoidosis through Toll-like receptor-2. Am J Respir Crit Care Med 2010; 181 (4):360–373.

37. Yakupova EI, Bobyleva LG, Vikhlyantsev IM, Bobylev AG. Congo Red and amyloids: history and relationship. Biosci Rep 2019; 39 (1):BSR20181415.

38. Benson MD, Berk JL, Dispenzieri A, Damy T, Gillmore JD, Hazenberg BP, et al. Tissue biopsy for the diagnosis of amyloidosis: experience from some centres. Amyloid 2021:1–6.

39. Howie AJ, Brewer DB, Howell D, Jones AP. Physical basis of colors seen in Congo red-stained amyloid in polarized light. Laboratory Investigation 2008; 88 (3):232–242.

40. Howie AJ, Brewer DB. Optical properties of amyloid stained by Congo red: history and mechanisms. Micron (Oxford, England : 1993) 2009; 40 (3):285–301.

41. Westermark P. Subcutaneous adipose tissue biopsy for amyloid protein studies. Methods in molecular biology (Clifton, NJ) 2012; 849:363–371.

42. Kyle RA. Amyloidosis: a convoluted story. British Journal of Haematology 2001; 114 (3):529–538.

43. Rohini K, Surekha Bhat M, Srikumar PS, Mahesh Kumar A. Assessment of Hematological Parameters in Pulmonary Tuberculosis Patients. Indian J Clin Biochem 2016; 31 (3):332–335.

44. Shafee M, Abbas F, Ashraf M, Alam Mengal M, Kakar N, Ahmad Z, et al. Hematological profile and risk factors associated with pulmonary tuberculosis patients in Quetta, Pakistan. Pak J Med Sci 2014; 30 (1):36–40.

45. Chedid C, Kokhreidze E, Tukvadze N, Banu S, Uddin MKM, Biswas S, et al. Association of baseline white blood cell counts with tuberculosis treatment outcome: a prospective multicentered cohort study. International Journal of Infectious Diseases 2020; 100:199–206.

46. Kahase D, Solomon A, Alemayehu M. Evaluation of Peripheral Blood Parameters of Pulmonary Tuberculosis Patients at St. Paul’s Hospital Millennium Medical College, Addis Ababa, Ethiopia: Comparative Study. Journal of blood medicine 2020; 11:115.

47. Malhotra P, Agarwal R, Awasthi A, Jindal SK, Srinivasan R. How long does it take for tuberculosis to cause secondary amyloidosis? European Journal of Internal Medicine 2005; 16 (6):437–439.

48. Dember LM. Amyloidosis-associated kidney disease. Journal of the American Society of Nephrology 2006; 17 (12):3458–3471.

49. Pires D, Marques J, Pombo JP, Carmo N, Bettencourt P, Neyrolles O, et al. Role of Cathepsins in Mycobacterium tuberculosis Survival in Human Macrophages. Scientific Reports 2016; 6 (1):32247.

50. De Buck M, Gouwy M, Wang JM, Van Snick J, Opdenakker G, Struyf S, et al. Structure and Expression of Different Serum Amyloid A (SAA) Variants and their Concentration-Dependent Functions During Host Insults. Curr Med Chem 2016; 23 (17):1725–1755.

51. Seto S, Morimoto K, Yoshida T, Hiramatsu M, Hijikata M, Nagata T, et al. Proteomic Profiling Reveals the Architecture of Granulomatous Lesions Caused by Tuberculosis and Mycobacterium avium Complex Lung Disease. Frontiers in Microbiology 2020; 10 (3081).

52. Van Gerven N, Van der Verren SE, Reiter DM, Remaut H. The Role of Functional Amyloids in Bacterial Virulence. J Mol Biol 2018; 430 (20):3657–3684.

53. Kaur G, Kaundal S, Kapoor S, Grimes JM, Huiskonen JT, Thakur KG. Mycobacterium tuberculosis CarD, an essential global transcriptional regulator forms amyloid-like fibrils. Scientific Reports 2018; 8 (1):10124.

54. Wang L, Maji SK, Sawaya MR, Eisenberg D, Riek R. Bacterial Inclusion Bodies Contain Amyloid-Like Structure. PLOS Biology 2008; 6 (8):e195.

55. Alteri CJ, Xicohténcatl-Cortes J, Hess S, Caballero-Olín G, Girón JA, Friedman RL. <em>Mycobacterium tuberculosis</em> produces pili during human infection. Proceedings of the National Academy of Sciences 2007; 104 (12):5145–5150.

56. Gaiser A-K, Bauer S, Ruez S, Holzmann K, Fändrich M, Syrovets T, et al. Serum Amyloid A1 Induces Classically Activated Macrophages: A Role for Enhanced Fibril Formation. Frontiers in Immunology 2021; 12 (2509).

57. Lundmark K, Vahdat Shariatpanahi A, Westermark GT. Depletion of spleen macrophages delays AA amyloid development: a study performed in the rapid mouse model of AA amyloidosis. PLoS One 2013; 8 (11):e79104.

58. Kluve-Beckerman B, Liepnieks JJ, Wang L, Benson MD. A Cell Culture System for the Study of Amyloid Pathogenesis: Amyloid Formation by Peritoneal Macrophages Cultured with Recombinant Serum Amyloid A. The American Journal of Pathology 1999; 155 (1):123–133.

59. Williams A, Orme IM, Jr. WRJ, McShane H, Mizrahi V, Orme IM. Animal Models of Tuberculosis: An Overview. Microbiology Spectrum 2016; 4 (4):4.4.09.

60. Bucsan AN, Mehra S, Khader SA, Kaushal D. The current state of animal models and genomic approaches towards identifying and validating molecular determinants of Mycobacterium tuberculosis infection and tuberculosis disease. Pathog Dis 2019; 77 (4):ftz037.

